## Supplementary Figures for "Near-term forecasting of Covid-19 cases and hospitalisations in Aotearoa New Zealand"

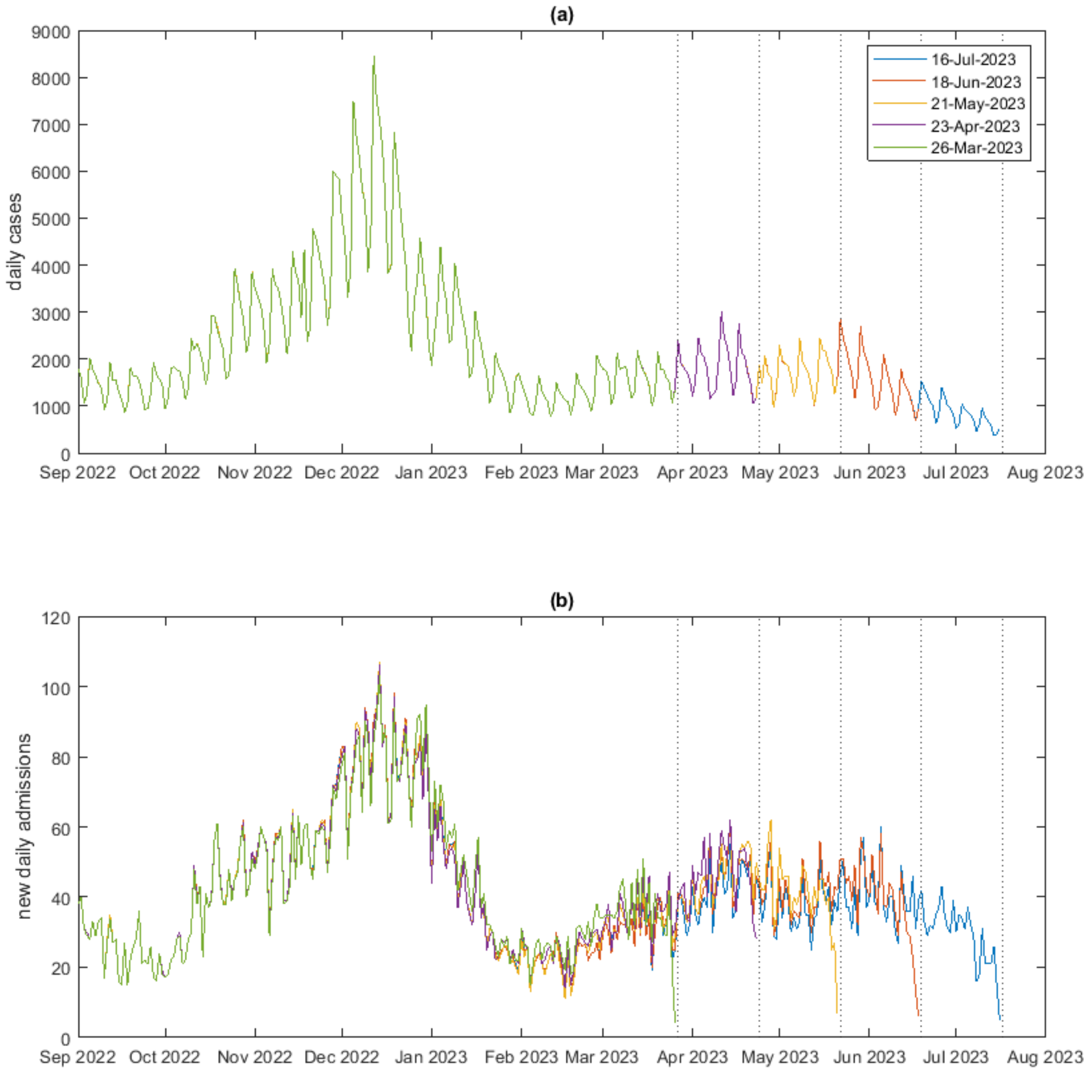


Figure S1: Data on (a) the number of daily reported cases and (b) the number of daily new hospital admissions, according to data supplied on 26 March 2023 (green), 23 April 2023 (purple), 21 May 2023 (yellow), 18 June 2023 (red) and 16 July 2023 (blue). There are negligible changes to the number of reported cases in subsequent datasets (note curves representing later datasets are largely hidden underneath curves for earlier datasets). However there are significant changes between datasets in the number of hospital admissions for a given day. In particular admissions for the most recent week of data are incomplete due to reporting lags. All forecasts produced in the paper are based on data that was available at the time of the forecast and are tested against the most recent available data.

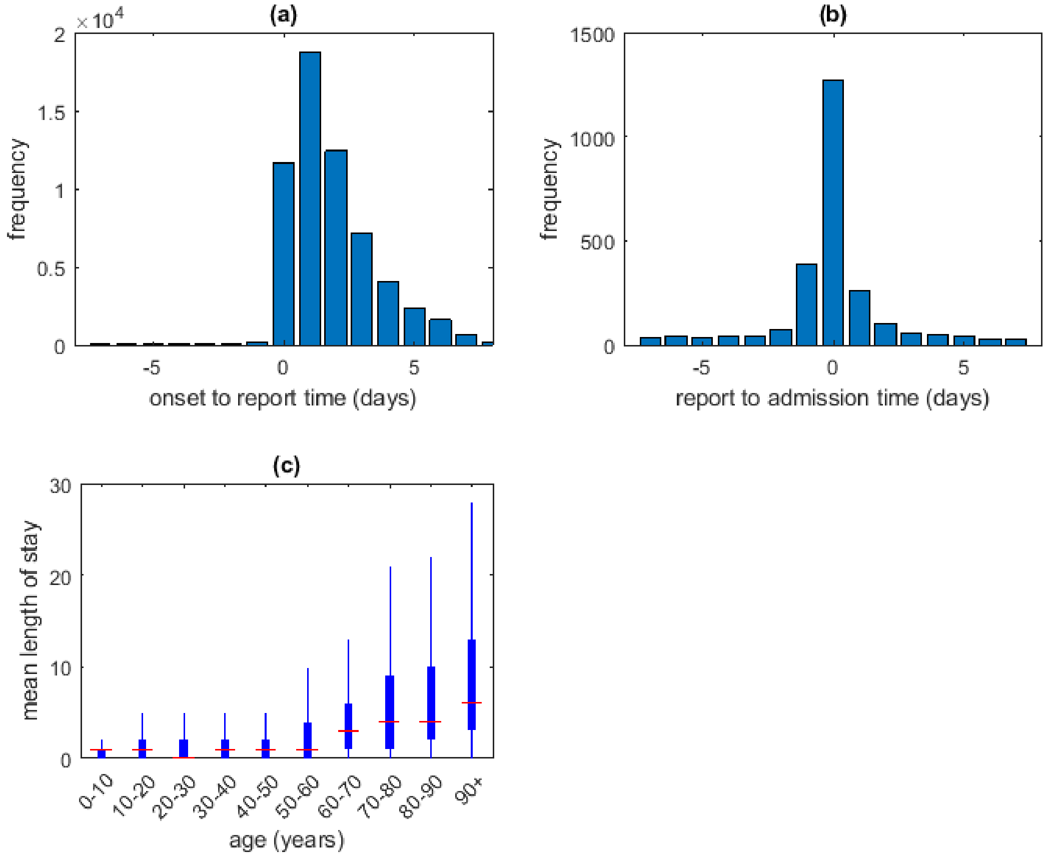


Figure S2: Empirically derived distributions of: (a) onset to report time (mean 1.9 days, s.d. 1.8 days, $n = 59292$); (b) report to admission time (mean 0.0 days, s.d. 2.1 days, $n = 2488$); (c) Covid-19-related length of hospital stay in 10-year age groups. Distributions calculated as described in Methods from the data as at 23 July 2023.

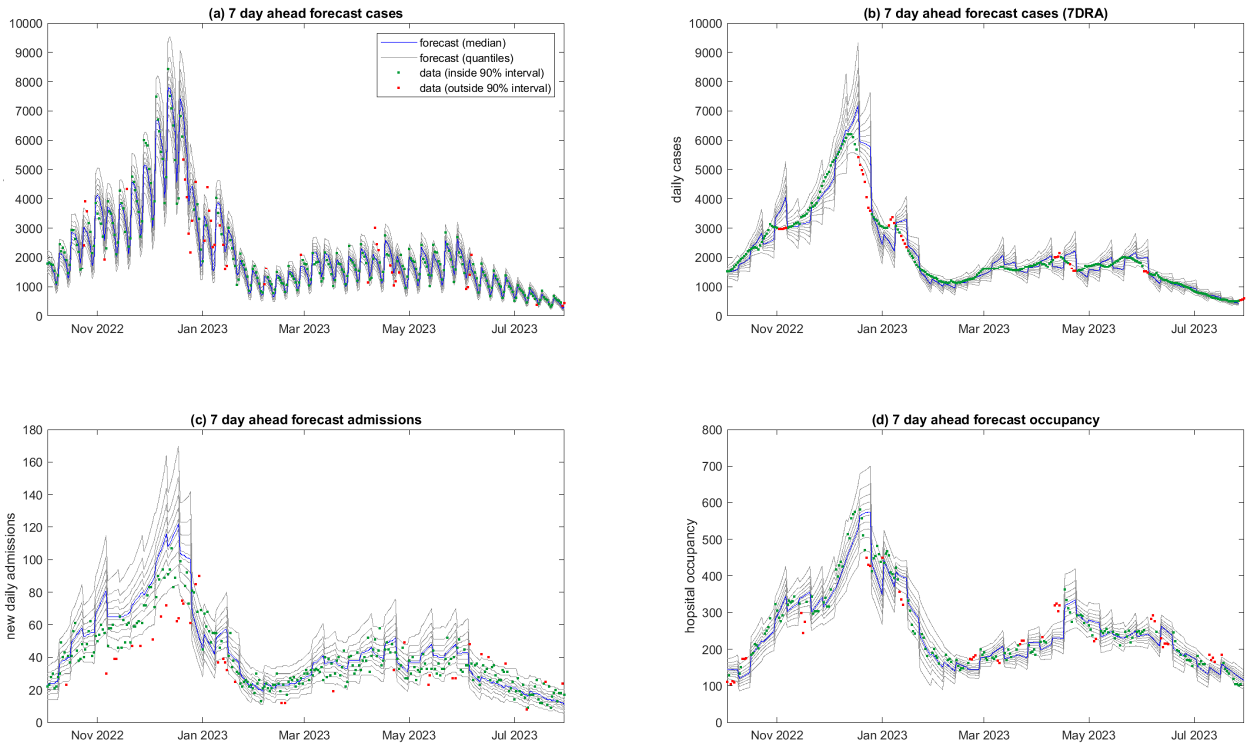


Figure S3: 7-day ahead forecast performance. Model results generated from data supplied at one of a series of weekly time points from 2 October 2022 to 23 July 2023, compared to actual data in the period 1-7 days subsequent to the date the data was supplied: (a) new daily cases; (b) smoothed daily cases (seven-day rolling average); (c) new daily hospital admissions; (d) hospital occupancy. Testing data was supplied on 20 August 2023 (i.e. 4 weeks subsequent to the last forecast). Weekly discontinuities in the forecasts are because each 7-day block represents a forecast generated from data supplied on a different date. Blue curve is the median and grey curves are the $5^{\text{th}}, 15^{\text{th}}, \ldots, 85^{\text{th}}, 95^{\text{th}}$ percentiles of $M = 10^5$ particles. Data points outside the $5^{\text{th}}$–$95^{\text{th}}$ percentile range of the forecast are shown in red.

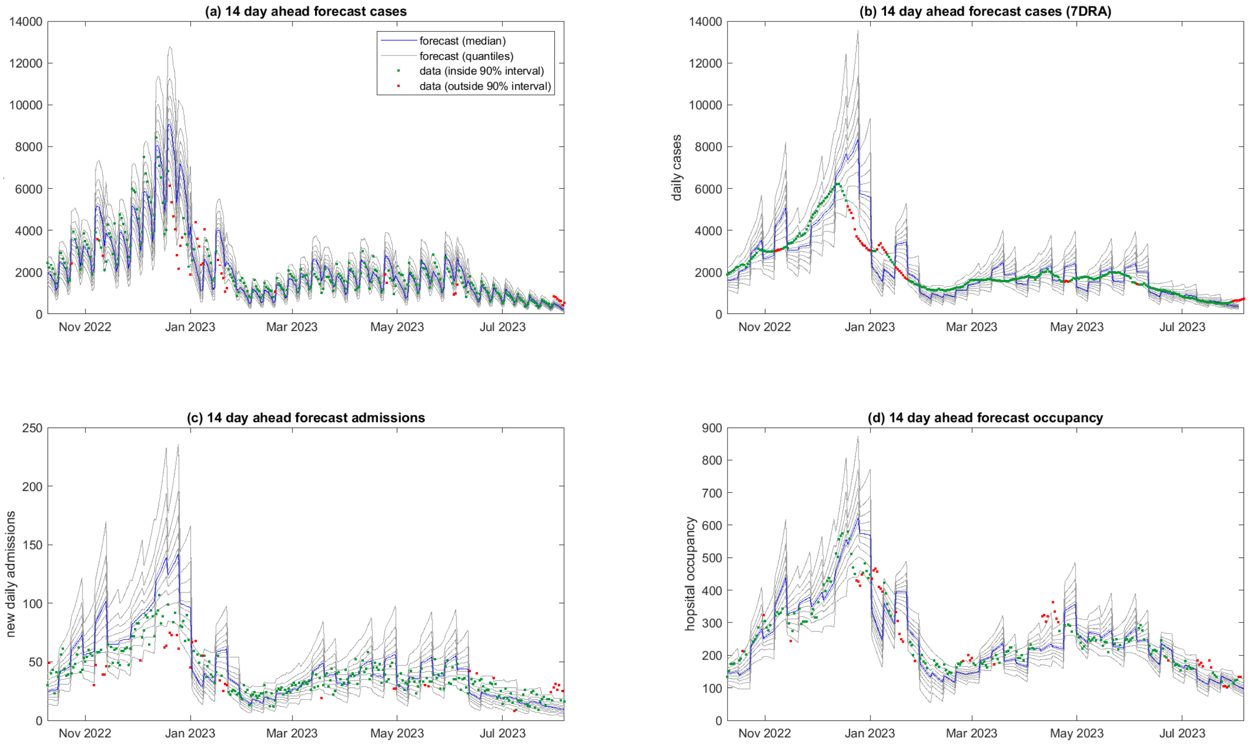


Figure S4: 14-day ahead forecast performance. Model results generated from data supplied at one of a series of weekly time points from 2 October 2022 to 23 July 2023, compared to actual data in the period 8-14 days subsequent to the date the data was supplied: (a) new daily cases; (b) smoothed daily cases (seven-day rolling average); (c) new daily hospital admissions; (d) hospital occupancy. Testing data was supplied on 20 August 2023 (i.e. 4 weeks subsequent to the last forecast). Weekly discontinuities in the forecasts are because each 7-day block represents a forecast generated from data supplied on a different date. Blue curve is the median and grey curves are the $5^{\text{th}}, 15^{\text{th}}, \ldots, 85^{\text{th}}, 95^{\text{th}}$ percentiles of $M = 10^5$ particles. Data points outside the $5^{\text{th}}$–$95^{\text{th}}$ percentile range of the forecast are shown in red.

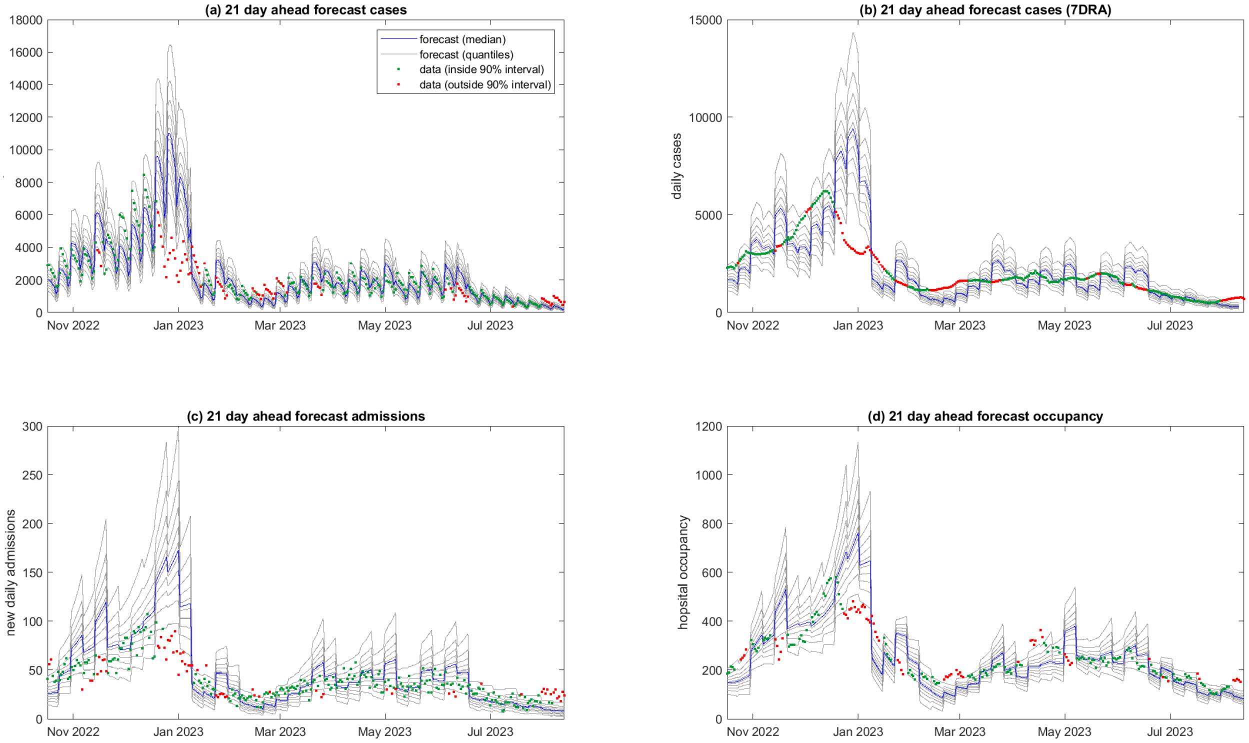


Figure S5: 21-day ahead forecast performance with a reduced value of the random walk standard deviation ($\sigma_R = 0.015$). Model results generated from data supplied at one of a series of weekly time points from 2 October 2022 to 23 July 2023, compared to actual data in the period 15-21 days subsequent to the date the data was supplied: (a) new daily cases; (b) smoothed daily cases (seven-day rolling average); (c) new daily hospital admissions; (d) hospital occupancy. Testing data was supplied on 20 August 2023 (i.e. 4 weeks subsequent to the last forecast). Weekly discontinuities in the forecasts are because each 7-day block represents a forecast generated from data supplied on a different date. Blue curve is the median and grey curves are the $5^{\text{th}}, 15^{\text{th}}, \ldots, 85^{\text{th}}, 95^{\text{th}}$ percentiles of $M = 10^5$ particles. Data points outside the $5^{\text{th}}$–$95^{\text{th}}$ percentile range of the forecast are shown in red.

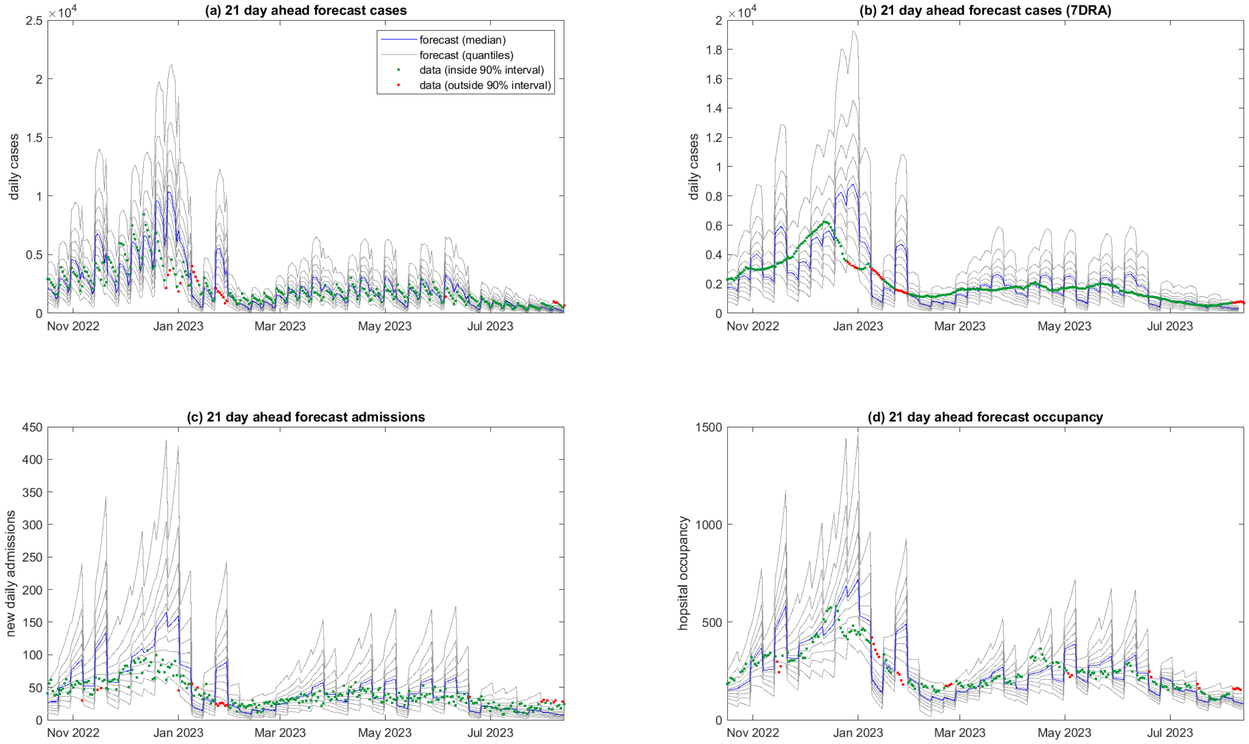


Figure S6: 21-day ahead forecast performance with an increased value of the random walk standard deviation ($\sigma_R = 0.035$). Model results generated from data supplied at one of a series of weekly time points from 2 October 2022 to 23 July 2023, compared to actual data in the period 15-21 days subsequent to the date the data was supplied: (a) new daily cases; (b) smoothed daily cases (seven-day rolling average); (c) new daily hospital admissions; (d) hospital occupancy. Testing data was supplied on 20 August 2023 (i.e. 4 weeks subsequent to the last forecast). Weekly discontinuities in the forecasts are because each 7-day block represents a forecast generated from data supplied on a different date. Blue curve is the median and grey curves are the $5^{\text{th}}, 15^{\text{th}}, \ldots, 85^{\text{th}}, 95^{\text{th}}$ percentiles of $M = 10^5$ particles. Data points outside the $5^{\text{th}}$–$95^{\text{th}}$ percentile range of the forecast are shown in red.

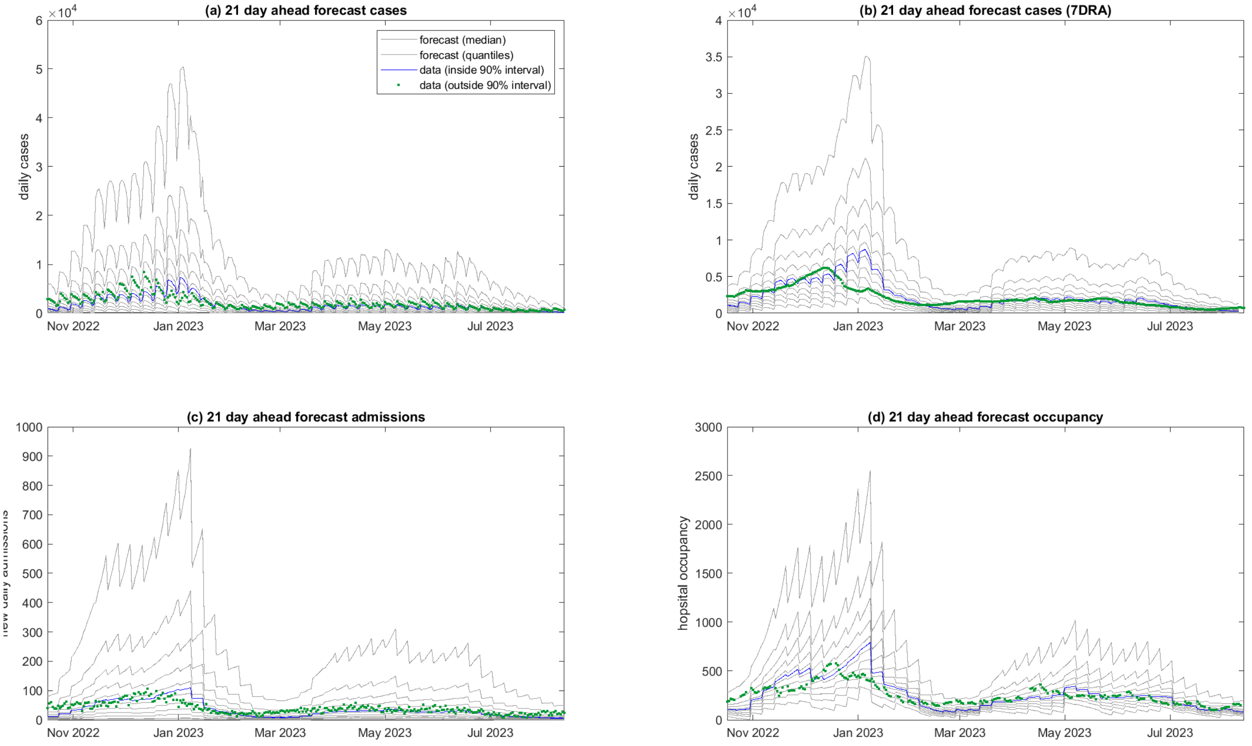


Figure S7: 21-day ahead forecast performance with increased variance in the model distributions for daily reported cases ($k_c = 1$) and daily admissions ($k_h = 1$). Model results generated from data supplied at one of a series of weekly time points from 2 October 2022 to 23 July 2023, compared to actual data in the period 15-21 days subsequent to the date the data was supplied: (a) new daily cases; (b) smoothed daily cases (seven-day rolling average); (c) new daily hospital admissions; (d) hospital occupancy. Testing data was supplied on 20 August 2023 (i.e. 4 weeks subsequent to the last forecast). Weekly discontinuities in the forecasts are because each 7-day block represents a forecast generated from data supplied on a different date. Blue curve is the median and grey curves are the $5^{\text{th}}, 15^{\text{th}}, \ldots, 85^{\text{th}}, 95^{\text{th}}$ percentiles of $M = 10^5$ particles. Data points outside the $5^{\text{th}}$–$95^{\text{th}}$ percentile range of the forecast are shown in red.

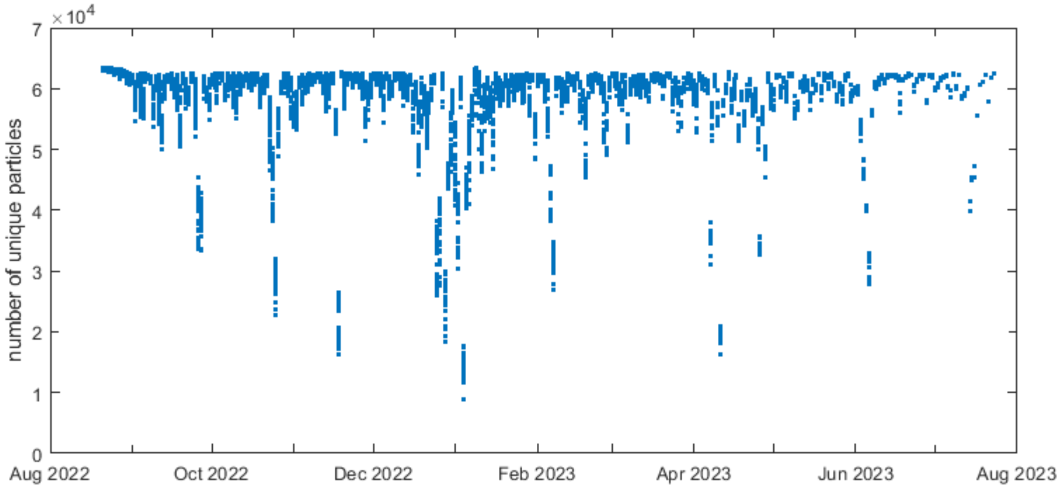


Figure S8: Number of unique particles in the bootstrap filter algorithm, showing pooled results across model runs for data supplied at one of a series of weekly time points from 2 October 2022 to 23 July 2023. The number of unique particles particles (out of $M = 10^5$ total particles) is almost always greater than $10^4$.
